# Consensus-based technical recommendations for clinical translation of renal Dynamic Contrast-Enhanced (DCE) MRI

**DOI:** 10.64898/2026.05.11.26352525

**Authors:** Ebony R. Gunwhy, Sila Kurugol, Suraj Serai, Aart J. van der Molen, Mohamed Abou El-Ghar, David L. Buckley, Paul D. Hockings, Richard A. Jones, Ruth P. Lim, Iosif A. Mendichovszky, Michael Pedersen, Hayley M. Reynolds, Luis C. Sanmiguel Serpa, Andrew Wentland, Frank G. Zöllner, Steven Sourbron, Ilona A. Dekkers

## Abstract

**Background:** Dynamic contrast-enhanced (DCE) MRI has the potential to be a useful tool for non-invasively assessing renal haemodynamics and function, however insufficient standardisation and difficulties in post-processing remain barriers to clinical translation.

**Purpose:** To develop expert consensus-based technical recommendations for performing renal DCE-MRI in humans, relating to aspects of patient preparation, MRI hardware and acquisition parameters, and data analysis.

**Study Type:** Systematic consensus process using an approximation to the two-step modified Delphi method.

**Population:** Not applicable.

**Field Strength / Sequence:** 1.5 T and 3 T / Renal gradient echo-based 3D DCE-MRI.

**Assessment:** An international panel of experts were recruited and surveyed following a modified Delphi method to create consensus-based technical recommendations. Key areas for consensus were initially identified through a mixture of online and in-person discussions, and an initial survey round consisting of open- and close-ended questions. Consensus statements were formulated and iteratively refined to create the final recommendations.

**Statistical Tests:** Consensus was defined as ≥ 75% agreement in response (excluding abstentions), and clear preference was defined as [60-74]% agreement among the experts. Statements with ≥40% abstentions were either excluded from subsequent survey rounds or recirculated as a modified statement.

**Results:** 22 experts initially participated in the Delphi panel, of which 16 responded to the first survey. 15 panellists responded to all subsequent surveys. Out of 46 statements, 37 reached consensus and one showed clear preference. ≥40% abstention was found in seven statements which were excluded from the final set of recommendations.

**Data conclusion:** These recommendations provide a starting point for MRI centres worldwide wishing to perform renal DCE-MRI, contributing to the harmonisation of DCE-MRI scan protocols and facilitating clinical translation. These recommendations provide a practical minimum technical dataset for renal DCE-MRI acquisition and analysis to improve cross-site comparability and support responsible clinical translation.

## Introduction

Dynamic contrast-enhanced magnetic resonance imaging (DCE-MRI) was initially developed to assess tissue perfusion and vascular permeability, often in oncology imaging in organs such as the brain and liver. Its use has since expanded to other organs, including the kidneys, where it is applied to evaluate renal perfusion and function in diagnosing and monitoring various adult and paediatric kidney disorders. DCE-MRI is a technique that involves the acquisition of sequential T_1_-weighted images following the administration of a gadolinium-based contrast agent (GBCA) to assess the dynamic passage of the agent through tissues (1). This enables the evaluation of tissue perfusion, vascular permeability, and overall organ function. DCE-MRI facilitates detailed assessment of kidney perfusion and filtration dynamics which are relevant for diagnosing and monitoring kidney diseases and evaluating the effects of therapeutic interventions (2).

Renal DCE-MRI can be used qualitatively for visually assessing the passage of the GBCA throughout the renal vasculature and parenchyma, followed by excretion into the collecting system over time. For example, delayed or reduced enhancement between the kidneys may be indicative of renal artery stenosis (3). Alternatively, symmetrically delayed enhancement or lack of excretion may be indicative of acute kidney injury (AKI) (4), and abnormal dilation of the collecting system is characteristic of (congenital) hydronephrosis (5).

Quantitative analysis has been evaluated for the estimation of functional parameters, such as (single-kidney) glomerular filtration rate (GFR), renal blood flow (RBF), and vascular permeability. While inulin is the reference standard and iohexol is increasingly used due to its practicality, DCE-MRI offers a more comprehensive assessment by combining functional estimation with high-resolution structural and haemodynamic data. Furthermore, while nuclear medicine techniques such as ⁹⁹ᵐTc-DTPA can provide split renal function, their utility is often limited in patients with severely impaired kidney function. In contrast, DCE-MRI can provide detailed, region-specific quantitative parameters even in complex anatomical or low-function scenarios.

Other 3D imaging techniques such as SPECT, CT and PET have been explored as alternative means for determining GFR, however MRI has the advantage of not requiring ionising radiation, and offers superior spatial resolution to molecular imaging techniques (5), facilitating acquisition of both functional and high resolution anatomical information in the same investigation. Arterial Spin Labelling (ASL) is another MRI technique which has shown promising clinical applications for measuring renal perfusion without the need for GBCAs (6). However, DCE-MRI offers advantages over ASL, such as increased spatial resolution and greater signal-to-noise ratio (SNR), coupled with the ability to assess GFR by using an exogenous agent that is freely filtered by the glomerulus (7).

The Cooperation in Science and Technology (COST) Action (CA16103) PARENCHIMA (8), which ended in 2021, has developed consensus-based technical recommendations for renal ASL (6), BOLD (9), DWI (10), T1 and T2 mapping (11), and Phase Contrast (12) MRI. This study, initiated by the Renal MR study group of the International Society for Magnetic Resonance in Medicine (ISMRM), aims to further contribute to these standardisation efforts for multiparametric renal MRI by providing expert, consensus-based technical recommendations for performing renal DCE-MRI to provide a basis for future research initiatives.

Despite the interest in renal DCE-MRI, research in this area remains more limited than for renal ASL. An important contributing factor is the requirement for GBCAs, which introduces additional operational and regulatory complexity, including adherence to established contrast safety frameworks such as the ACR Manual on Contrast Media (https://www.acr.org/Clinical-Resources/Clinical-Tools-and-Reference/Contrast-Manual) and the Contrast Media Safety Committee of the European Society of Urogenital Radiology guidelines (https://www.esur.org/esur-guidelines-2025/). In recognition of these considerations, the technical recommendations presented in this manuscript are accompanied by a dedicated overview of GBCA-related safety aspects relevant to renal MRI research. This complementary statement, prepared by members of the ISMRM MR Contrast Agents study group (AJM) and CMSC-ESUR working group (AJM, IAD) with expertise in contrast media safety, summarises the principal risks and precautions associated with GBCA use in renal imaging, with the aim of supporting safe and consistent practice across institutions.

### Overview of technical parameters

#### Relationship between DCE-MRI signal and GBCA concentration

DCE-MRI is acquired using a T_1_-weighted sequence to capture serial MR images over time. After an initial baseline scanning period (pre-contrast), a bolus of gadolinium-based contrast agent (GBCA) is administered intravenously to the patient. To ensure rapid and standardised delivery of the GBCA, a power injector can be used for GBCA administration, followed by a saline flush injected at the same rate (13,14). The resulting MR images provide a measurement of longitudinal relaxation time (T_1_) changes in tissue over time, providing a means for observing the passage and tissue accumulation of GBCA, which is reflected as signal enhancement due to T_1_ shortening.

Signal enhancement is first observed in the renal artery, reflecting bolus arrival of the GBCA, followed by uptake in the renal vasculature and the tubular system, and then clearance as the agent is evacuated from these spaces (***Fig. 1***).

**Fig. 1.**
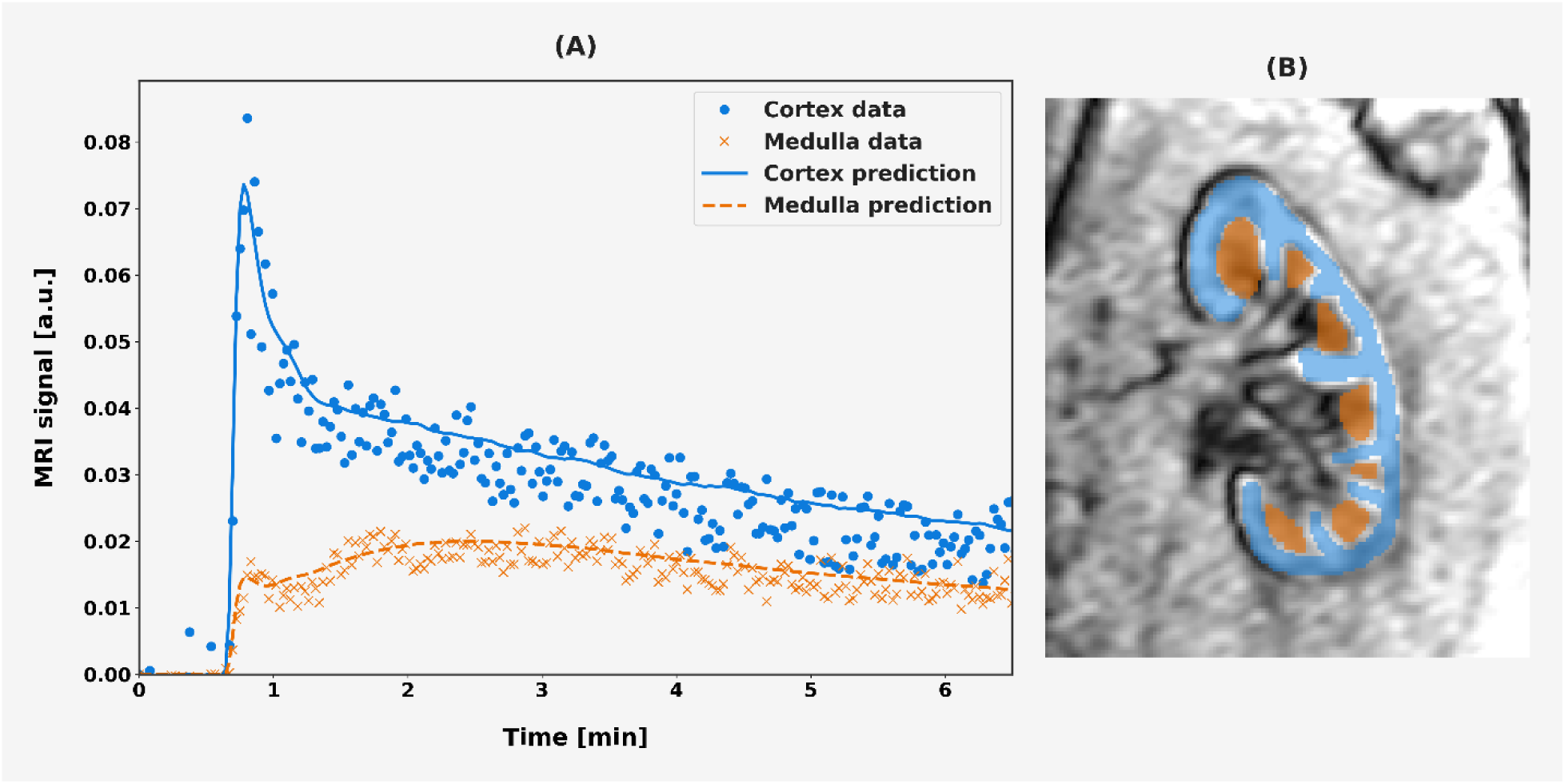
DCE-MRI data provided by the Prognostic imaging biomarkers for diabetic kidney disease (iBEAt) study (16) and analysed with the ‘KidneyCortMed’ model from dcmri.org (33), for illustration purposes only. **(A)** Example DCE-MRI signal intensity plots demonstrating gadolinium based contrast agent (GBCA) dynamics within renal cortex and medulla regions of interest (ROIs). Data was acquired with a spoiled gradient echo (SPGR) MRI sequence at a field strength of 3 T. Plot shows observed data overlaid with predicted data after fitting to a SPGR signal model. a.u. = arbitrary units. **(B)** Renal cortex and medulla ROIs segmented upon the DCE-MR image. An average ROI pixel intensity was obtained at each timepoint and used to the plot the data in (A). ROI colours match those shown in the legend in (A).

For quantitative analysis of DCE-MRI, a method for deriving the GBCA concentration from the observed MRI signal intensity is required. As the change in tissue relaxation rate, R1 (= 1/T1), is proportional to the GBCA concentration, an analytical expression can be used to describe the relationship between the signal intensity and R1 (15), which is dependent on the MRI sequence and sequence parameters (e.g., TR, TE, FA). Since the kidney is highly perfused, GBCA doses lower than the full standard clinical dose of 0.1 mmol/kg of body weight are sometimes administered (16). This also helps to reduce such artefacts as T * effects within the renal vessels (13,17,18).

#### Renal DCE-MRI planning and sequence parameters

Experimental variables such as patient hydration status, diet, and medication intake should be carefully considered prior to renal DCE-MRI acquisition. Standardisation is not always possible; however, all these factors can potentially affect results, therefore recording these factors prior to scanning may aid in understanding potential variation.

DCE-MRI is typically acquired with a spoiled gradient echo (SPGR) MRI sequence at field strengths of 1.5 T or 3 T. DCE-MRI generally does not benefit from higher field strengths, though these are often used because DCE-MRI is applied together with other scans that benefit from increased SNR. However, this comes with an increased risk of artifacts, B_1_ magnetic field inhomogeneity, and specific absorption rate (SAR) constraints (19).

To reduce artifacts introduced from patient respiratory motion, multiple breath-hold techniques may be employed. However, this can be difficult for some patients and variability can be encountered between breath-holds. Alternatively, free breathing approaches may also be used, followed by post-processing image registration to correct respiratory motion effects (20).

The choice of MRI sequence parameters for renal DCE-MRI is dependent on whether the analysis planned is quantitative or qualitative. For example, both 2D and 3D readout schemes can be used to acquire renal DCE-MRI, however 3D MRI offers advantages for quantitative analysis as it typically provides superior through-plane spatial resolution, signal-to-noise ratio, and greater anatomical coverage (21). Similarly, quantitative analysis may benefit from faster temporal resolution to more accurately capture the rapid renal uptake and washout of the GBCA (22). A smaller slice thickness and in-plane pixel size may also help to reduce partial volume effects, albeit at the expense of reduced SNR and longer acquisition times.

A pre-contrast calibration T_1_-mapping sequence is also beneficial for more accurate quantitative analysis. Various techniques are used for this, such as multiple inversion recovery (IR) times or variable flip angles (vFA) (13). To reduce errors in T_1_ measurements occurring as a result of magnetic field inhomogeneities, B_0_ and B_1_-mapping should also be considered (11).

A short TR and TE is typically required for renal DCE-MRI, for both quantitative and qualitative applications.

#### Renal tracer kinetic modelling

The uptake of GBCA can be assessed both qualitatively, from visual inspection of the images, and quantitatively. For quantitative analysis, a tracer kinetic model is needed to demonstrate how the GBCA is expected to pass through a tissue space. Various models have been proposed for this, modelling the nephron as a chain of compartments (23).

Using too few compartments to model GBCA flow through the renal system can oversimplify GBCA dynamics and result in underfitting of the DCE-MRI data. Increasing the number of nephron compartments can result in a more realistic physiological model and the estimation of a greater number of parameters. However, increasing complexity can also result in overfitting of the DCE-MRI data.

Variations of the two-compartment models are most frequently cited (24–29).

To account for tubular outflow, a two-compartment filtration model (2CFM) (28,29) has been proposed which allows for the quantification of both renal perfusion and filtration. In this model, GBCA is transported from an arterial compartment via Plasma Flow (F_p_ [mL/min/100 mL]) into a glomerular plasma compartment. A fraction of GBCA from the glomerular plasma compartment is then filtered into a tubular compartment via Tubular Flow (F_t_ [mL/min/100 mL]). The Glomerular Filtration Rate (GFR [mL/min/1.73 m^2^]) can then be calculated by multiplying the F_t_ with the volume of both kidneys and scaling with body surface area.

To use these models in practice, renal regions of interest (ROIs) placed upon the DCE-MR image and feeding artery are required. These can be ROIs of the whole kidney, cortex, or cortex and medulla when using cortico-medullary modelling. A measurement of the arterial input function (AIF) of GBCA concentration into the renal compartment is also required. From a modelling perspective, this would ideally be measured in the renal artery at the most direct point of entry into the renal compartment.

However, this is difficult to achieve accurately in practice due to the minimal voxel size possible in DCE-MRI being greater than, or on the order of the size of the renal arterial diameter, which risks partial volume artifacts in the MRI signal. Consequently, the AIF is typically measured in the larger, descending aorta, or from a reference organ, such as the spleen, or by using a population- (30,31) or model- based AIF (32).

### Safety aspects related to the use of GBCA in renal imaging

Risks historically associated with GBCAs are well documented and have evolved in their interpretation over time, encompassing hypersensitivity reactions as well as concerns specific to patients with impaired kidney function, most notably NSF and contrast-induced acute kidney injury (CI-AKI). Over the past decades, and particularly with the transition from linear to more stable macrocyclic GBCAs, the absolute risks of these complications have declined substantially, resulting in a markedly improved overall safety profile.

With regard to the use of GBCA, the most important safety aspect is the infrequent occurrence of immediate (<1 h) or non-immediate (1h - 8 weeks) hypersensitivity reactions. Large studies in adult patient cohorts have shown a very low incidence of 0.06-0.17%, with severe reactions occurring in only 0.003-0.006% (34,35). Main risk factors for a (recurrent) hypersensitivity reaction include prior hypersensitivity to GBCA and history of (acute) allergic diseases. Recently, updated multidisciplinary guidelines for management of hypersensitivity reactions (36), or prevention of recurrent hypersensitivity reactions (37–39) have been published by ACR, ESUR and CAR.

NSF is a severe fibrosing disorder associated with the use of linear GBCAs in patients with poor renal function (estimated GFR < 30 ml/min/1.73m^2^ or dialysis) (40). The condition received substantial clinical and media attention when it was initially recognised. With the transition to more stable macrocyclic agents (current preferential use of macrocyclic GBCA in the European Union; ACR Group II GBCA in USA), however, instances of NSF in contemporary practice have become rare (41–43). Large post-marketing surveillance studies have reported no unconfounded cases of NSF associated with macrocyclic GBCAs (44,45).

For iodine-based contrast media used in CT, there has historically been strong emphasis on preventing CI-AKI (i.e. acute kidney injury occurring within 48h after contrast administration), particularly in patients with reduced renal function. For GBCAs, a clear association with CI-AKI has not been established. Given the 5-10 times lower molar dose of GBCA compared to iodine-based contrast media, and the low osmolar load of GBCA administration, CI-AKI after GBCA administration is unlikely when approved doses are administered (46,47), even in children (48). As a precaution, in clinical practice GBCAs are administered to patients with an eGFR < 30 mL/min/1.73 m² only after careful weighing of individual risks and expected diagnostic benefit.

Small amounts of GBCA retention in the human body have been reported after repeated administrations. Although >99.9% of the dose is renally excreted, a minor fraction enters a slowly exchanging compartment, leading to low-level, transient retention even with macrocyclic agents (49–51). These traces gradually clear, and no clinical symptoms have been identified in association with gadolinium retention over more than a decade of investigation (52).

In paediatric imaging, additional caution is warranted due to longer life expectancy and the potential need for repeated examinations; therefore, the use of the lowest effective dose of a macrocyclic gadolinium-based contrast agent is recommended. Importantly, contemporary evidence indicates no increased risk of acute kidney injury or clinically relevant adverse outcomes following contrast-enhanced MRI in children, including those with normal or mildly impaired renal function, when current safety guidelines are followed (48,52).

## Materials and Methods

Recruitment for the consensus panel began in August 2022, with an initial meeting conducted virtually to provide information on the initiative and to garner interest from a broad range of experts. Experts with publication records and/or research interests in renal DCE-MRI were identified and invited to participate. Panellist experience was intended to be as varied and representative as possible, with invitations extended globally to experts from multiple institutions and from both clinical and non-clinical fields. Delphi survey respondents who consented to authorship are included as co-authors of this manuscript.

For developing the consensus statements, an approximation to the two-step modified Delphi method (53,54) as described in the founding paper of the initial PARENCHIMA consensus initiative (8), was used. This involved iterative survey rounds, circulated electronically to the panel via Google Forms (<u>Supplementary material, ‘Iterative survey rounds.zip’</u>), where proposed consensus statements were modified at each iteration based on responses received during the previous round.

An overview of the project timeline is provided in ***Fig. 2***. Panellists were initially assigned to three working groups (A. participant preparation and hardware considerations; B. sequence parameters, including motion correction; C. data analysis and reporting) and asked to draft a list of topics relevant to renal DCE-MRI which should be considered when developing the statements. These were then combined with findings from literature and statements from previous renal MRI consensus work and formulated by the panel co-chairs (E.R.G. and I.D.) into a survey for the first iteration of the Delphi process (June 2023). This first survey round consisted of a mixture of 38 closed- and open-ended questions, with participants encouraged to provide detailed responses based on their experiences and expertise. Results of the first electronic survey were discussed in-person at the Fifth International Renal Imaging Meeting in Ghent, Belgium (September 10-11, 2023).

**Fig. 2.**
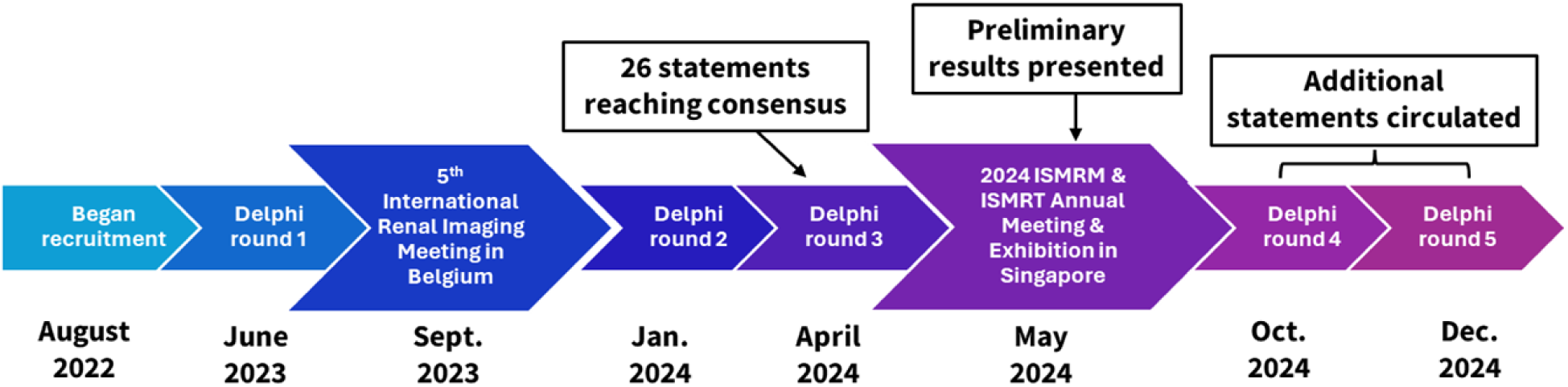
An overview of the renal DCE-MRI consensus project timeline.

Based on these discussions and responses from the first survey, a follow-up survey was constructed (second round). This second survey was formulated as a set of 38 ‘consensus statements’ circulated to the panel (January 2024). Five statements were circulated in a third survey round (April 2024) consisting of four statements reformulated from the previous round and one new statement.

Preliminary results from survey rounds 1-3 were presented at the ISMRM & ISMRT Annual Meeting & Exhibition in Singapore (May 2024) (55), with a view to promoting further discussion at the meeting and widening expert participation. Based on feedback, 17 additional statements relating to technical, sequence-specific MRI parameters and analysis were proposed and circulated to the panel in two subsequent survey rounds (October and December 2024). For these two final rounds, panellists were asked to provide responses to statements taking into consideration the statements which had already reached consensus from previous rounds, i.e., based on MRI protocols which incorporate the parameters that had already been defined in the preliminary set of recommendations.

For each statement in survey rounds 2-5, panellists were asked to specify a single response of ‘Strongly Agree’, ‘Agree’, ‘Neutral’, ‘Disagree’, or ‘Strongly Disagree’. Panellists were instructed to select ‘Neutral’ for statements where they did not consider themselves to be sufficiently knowledgeable to answer the question. In addition, panellists were always provided with a free-text option to elaborate on their response. As in previous work, a “traffic light” system was used to determine the statements included for consensus (see ***Fig. 3***). Statements were required to achieve “green light” status (≥ 75% agreement) to reach consensus. “Orange light” status ([60-74]% agreement) was assigned to statements which did not reach consensus yet indicated a clear preference among the experts. “Red light” status ([50-59]% agreement) was assigned to open issues, where no clear preference was indicated among the experts.

**Fig. 3.**
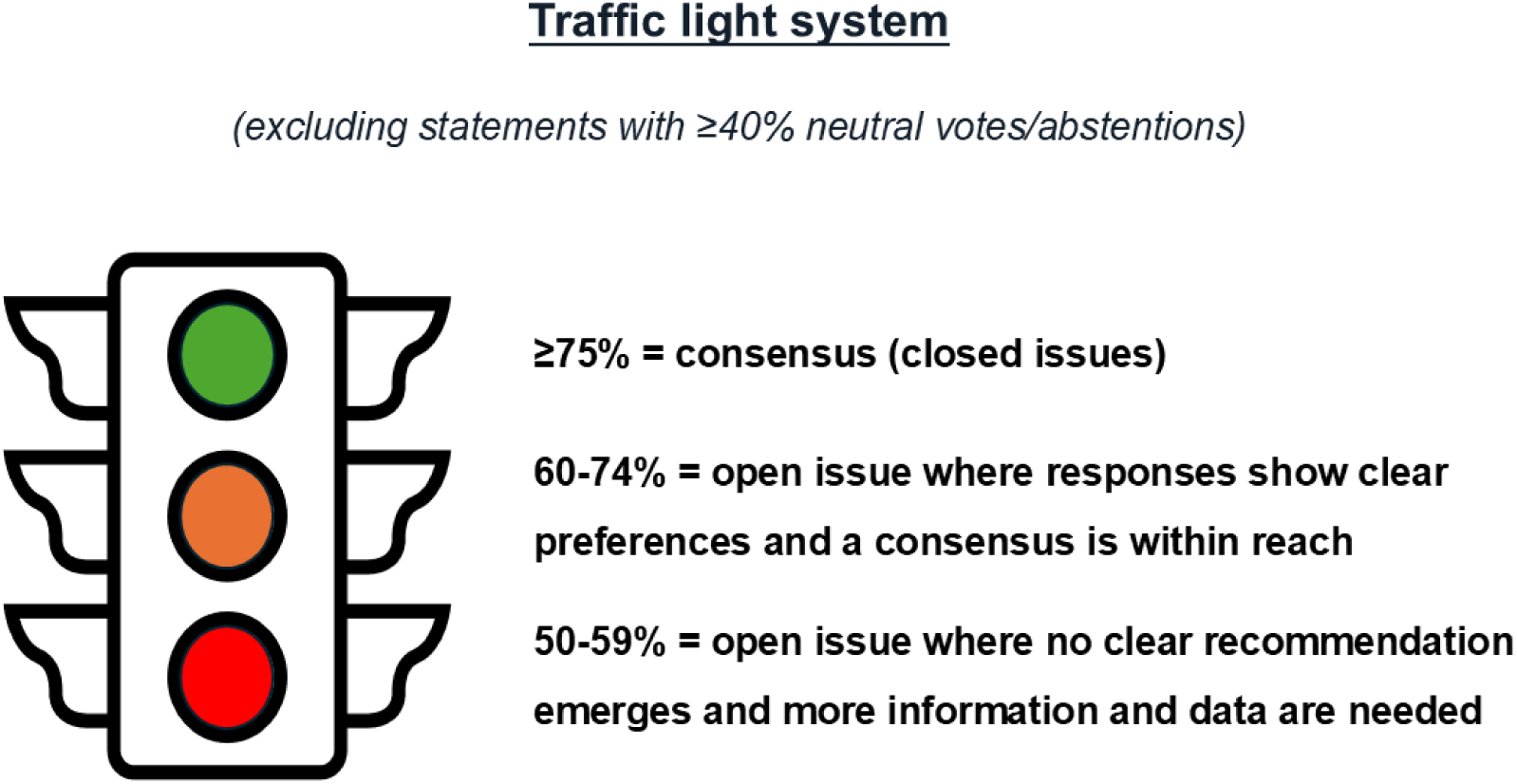
Schematic of the “traffic light” system coding used to define consensus recommendations.

Abstentions were excluded from the calculation of consensus percentages. Statements with ≥40% abstentions were either excluded from subsequent survey rounds or recirculated after modifying the statement.

## Results

In total, 22 experts participated in the initial Delphi panel meeting, of which 16 responded to the first survey. ***Fig. 4*** shows the countries of residence of each of the expert panel members who participated in the first survey. A total of 15 panellists responded to all subsequent surveys, as one respondent felt they no longer had relevant experience to continue participating. This panel size falls within the range of eight to 23 participants suggested for Delphi panels in health sciences research (56). Of the 15 experts who responded to all survey rounds, ten had a background in physics, seven in clinical radiology, four in computing, and three in engineering disciplines. Most experts were found to have experience in using renal DCE-MRI for quantitative research applications in adult populations, with only 26.67% of respondents using renal DCE-MRI in both clinical and research settings. However, routine use in paediatric studies for clinical purposes was also reported. Populations with whom respondents reported having experience using renal DCE-MRI included patients with chronic kidney disease, diabetes, congenital kidney abnormalities, and renal cell carcinoma, healthy volunteers, paediatric patients, and kidney transplant recipients, as well as animal models. A summary of panellist background and experience in renal DCE-MRI is provided in ***Fig. 5***.

**Fig. 4.**
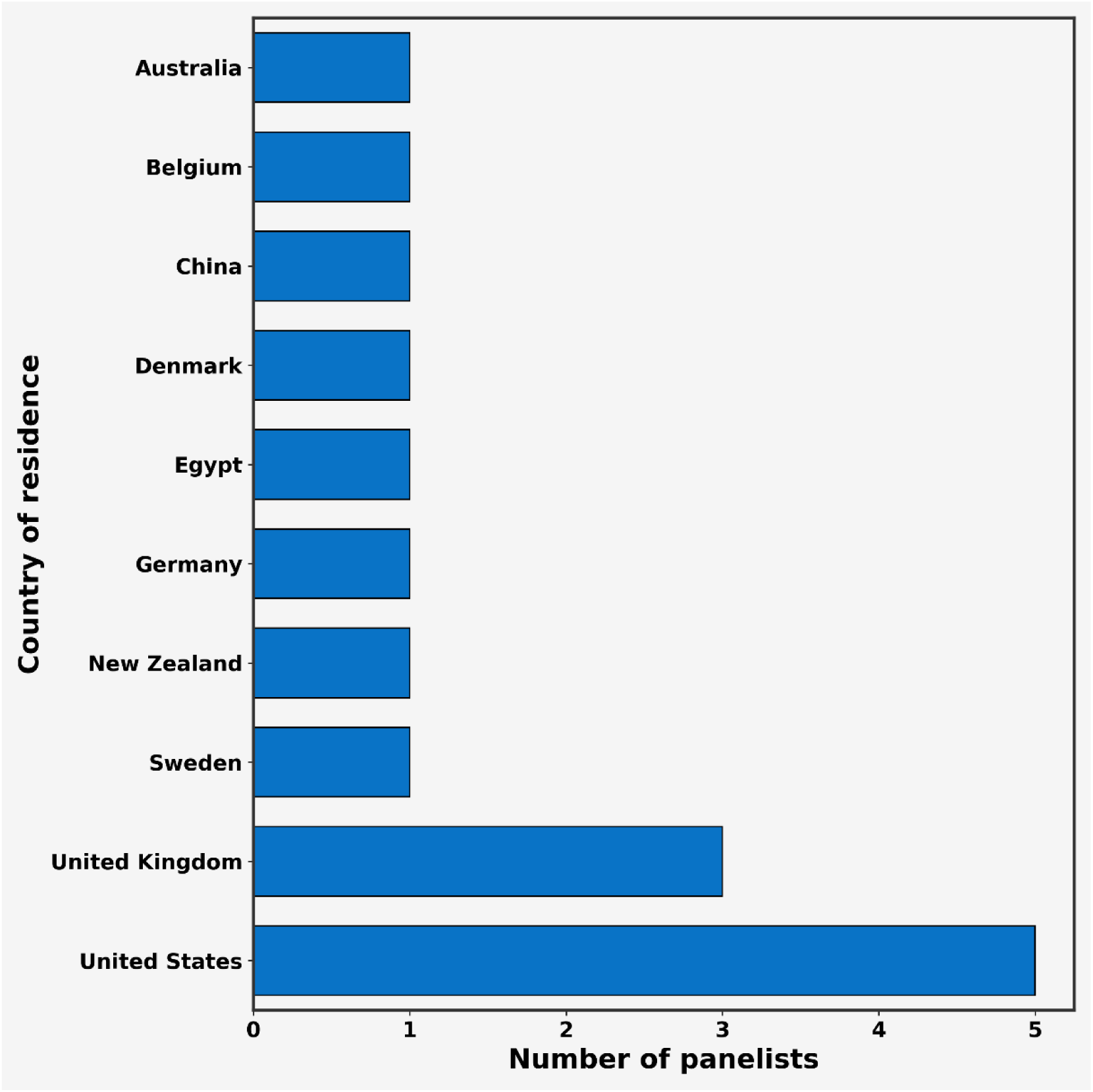
A summary of the countries of residence of the renal DCE-MRI expert panel members that responded to the first Delphi survey.

**Fig. 5.**
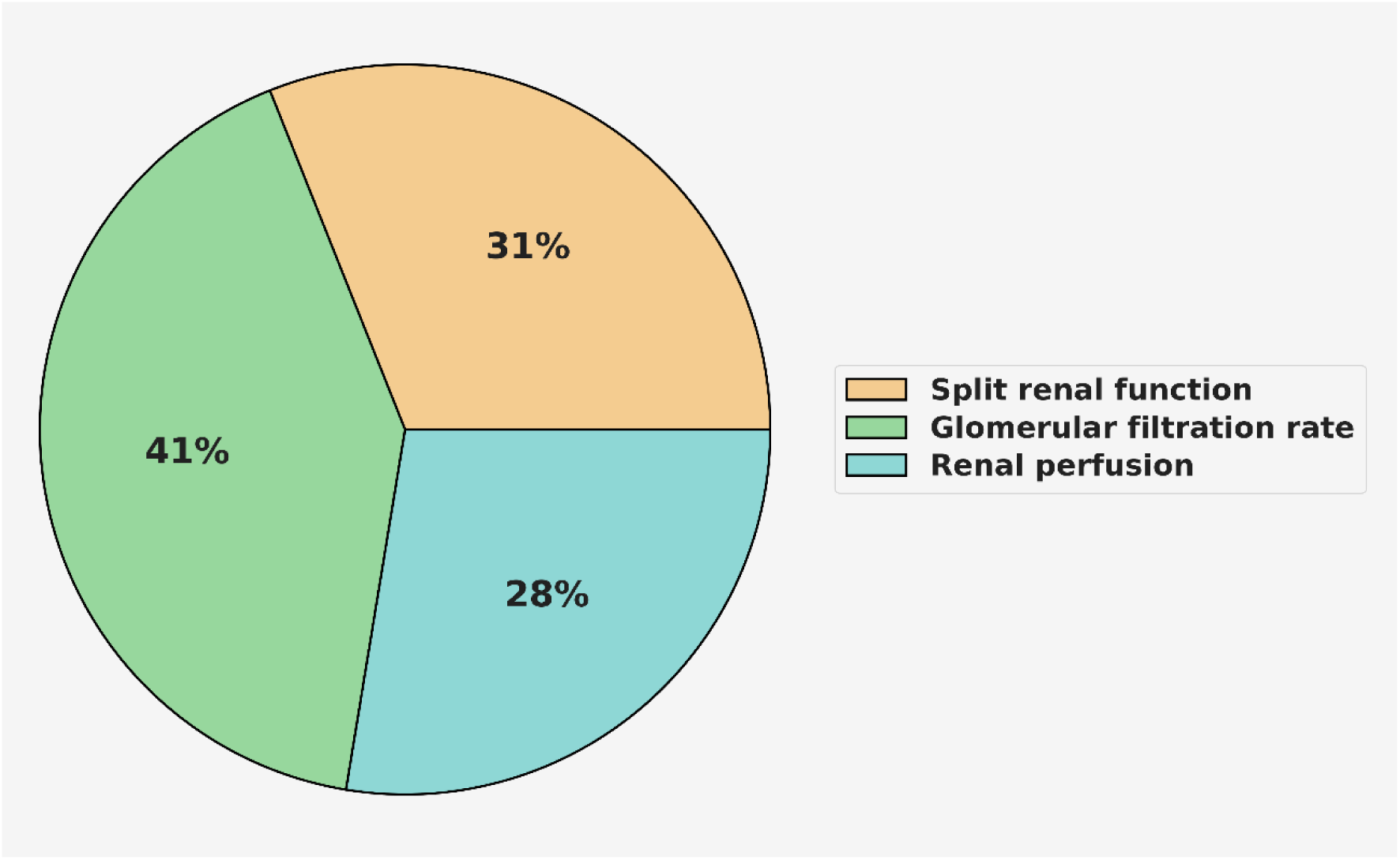
A summary of renal DCE-MRI quantitative use cases reported by the expert panel.

During the second survey round, 22 statements achieved immediate consensus. 11 statements which did not reach consensus were reformulated into four new statements. The remaining five statements which did not reach consensus were excluded from subsequent rounds. During the third survey round, consensus was achieved in three out of the four reformulated statements, and in one additional new statement, resulting in the inclusion of four additional consensus recommendations. The remaining statement which did not reach consensus was excluded from subsequent rounds.

During the fourth survey round, one statement reached immediate consensus and was included in the final recommendations. Three statements did not reach consensus and were excluded. The remaining 13 statements were reformulated into ten new statements. In the fifth and final round, all ten of the reformulated statements achieved consensus.

This resulted in a total of 37 consensus statements included in the final set of recommendations (Supplementary material, Table 2). Of these, 100% agreement was reached in 23 (∼62%) statements. In total, nine statements did not achieve consensus and were completely excluded (i.e., not reformulated) from the final set of recommendations (Supplementary material, Table 3).

Overall, for the statements reaching consensus, the average agreement and abstention levels were 95.3 ± 7.1% and 13.9 ± 10.6%, respectively.

### Final recommendations

Consensus-based recommendations for a renal DCE-MRI protocol are summarised in **Table 1**.

**Table 1.** Consensus-based recommendations for a renal DCE-MRI protocol.

| **Renal DCE-MRI Protocol** |  |
| --- | --- |
| *Patient preparation* | • Normal hydration status (when clinically appropriate)<br>• Record hydration status, dietary intake, and medication use |
| *Gadolinium-Based Contrast Agents (GBCA)* | • <0.1 mmol/kg of body weight<br>• Injected with a power injector<br>• Followed by saline flush at same rate of GBCA injection<br>• Administered after a sufficient number of baseline volumes ($\geq 5^*$) have been recorded |
| *Field Strength* | 1.5 T or 3 T |
| *Coils* | Body coil transmitter; multi-channel receiver coil |
| *Patient positioning* | Supine; kidneys in isocentre of magnet |
| *Preparatory sequence* | T1-mapping |
| *Sequence* | 3D gradient echo-based, |
| *Readout* | Cartesian or radial |
| *Orientation* | Coronal; coronal oblique |
| *Field of view* | Include both kidneys and major vessels; minimise aorta inflow artifacts |
| *Breathing mode* | Free breathing |
| *Flip angle (FA)* | 10-13 degrees, set **before** Repetition Time |
| *Repetition Time (TR)* | Shortest available for the given FA |
| *Echo Time (TE)* | Shortest available for the given FA and TR |
| *Slice Thickness* | 1-3 mm |
| *In-plane acquired pixel size* | $\leq 3$ mm |
| *Temporal resolution* | Shortest possible, ideally $\leq 3$ seconds |
| *Scan duration* | Up to 7 minutes |
| *Motion correction* | Post-processing image registration |
| *Arterial Input Function (AIF)* | Abdominal aorta if possible, otherwise replace with population- or model- based AIF |
| *Renal compartment model* | Two-compartment (glomerular plasma and tubular fluid) |
| *Units* | Glomerular Filtration Rate (GFR) in [mL/min/1.73 m<sup>2</sup>]; Tubular Flow (F<sub>t</sub>) and Plasma Flow (F<sub>p</sub>) in [mL/min/100 mL] |
| *Reporting* | Mean and standard deviation (when measuring pixel-based parameters) |

\*QIBA Dynamic Contrast Enhanced (DCE-MRI) Biomarker Committee: DCE-MRI V.2, Consensus Profile. 2023.

#### Recommendations regarding patient preparation (n = 4)

In the final recommendations, experts came to the consensus that dietary intake may influence functional measurements of renal DCE-MRI, and therefore dietary control should be considered prior to scanning. Experts also agreed that patients should be scanned in normal hydration status when clinically appropriate. However, experts expressed that this may be difficult in some populations, and responses were varied regarding the specific dietary or hydration status management required. For example, some experts advised that fluid intake should be avoided prior to scanning, while others advised that it was important for patients to be well hydrated, and in some cases should be provided additional hydration. Expert advice regarding dietary intake ranged from light meals reduced in protein to fasting prior to scanning.

Similarly, expert advice regarding specific patient medication use prior to scanning was varied, as this is dependent upon the population being scanned and the aim of the renal DCE-MRI assessment. Therefore, rather than providing a recommendation on standard dietary, hydration, and medication requirements, it was agreed by consensus that patient dietary intake, hydration status, and medication use should be recorded prior to renal DCE-MRI scanning.

#### Recommendations regarding renal DCE-MRI hardware (n = 3)

Experts reported experience with a variety of vendors: Siemens (n = 12), GE (n = 4), Philips (n = 6). By consensus, experts agreed that either 1.5 T or 3 T scanners could be used for renal DCE-MRI scanning, along with a body transmitter and multi-channel receiver coil. No consensus was found regarding the specific number of channels that should be used in the receiver coil, however clear preference (∼73% agreement) was found when experts were asked to respond to the statement ‘the coils should be adjusted to match the size of the patient’. Some experts highlighted that hardware choices were limited based on what was available at their centre, therefore they could only advise based on what was available to them.

Specific advice regarding contrast pump settings was varied, however it was agreed by consensus that contrast agents must be injected with a power injector rather than manually.

#### Recommendations regarding use of gadolinium-based contrast agents (n=4)

Experts agreed by consensus that GBCA doses administered should be body weight dependent and lower than the full standard clinical dose (in general <0.1 mmol/kg of body weight). Some experts commented that they used low GBCA doses to ensure concentration and signal intensities remained within the linear range. It was also agreed that injection should occur after a sufficient number of baseline volumes (≥5 (57)) have been recorded and a saline flush should be used at the same rate of injection. In general, it can be noted that bolus timing and injection rate (e.g., via power injector, usually 1–2 mL/s followed by saline flush) should be taken into account when administering GBCA.

#### Recommendations regarding renal DCE-MRI planning and sequence parameters (n = 19)

In the final recommendations, it was agreed by consensus that patients should be scanned in supine positioning and instructed to breathe freely for the duration of the acquisition. Experts commented that measures should be taken to ensure comfort of the subject, for example entering feet-first may minimise the risk of claustrophobia. It was also noted that in some cases, a breath-hold may be required depending on the specific sequence used. However, overall consensus was not agreed upon regarding these matters.

By consensus, it was agreed that coronal or coronal oblique orientations, and a field of view (FOV) which includes both kidneys positioned within the isocentre of the magnet, and extending to include major vessels are recommended for obtaining renal DCE-MRI. However, it was also recommended that the FOV should be positioned carefully (by centring it around the kidneys in the superior-inferior axis) to minimise inflow artifacts in the aorta. Regarding slice thickness and in-plane acquired pixel size, consensus for specific values covering a range of use cases was not found. However, recommendations for ranges of values for measurement of perfusion specifically were agreed by consensus (slice thickness: 1 - 3 mm; in-plane acquired pixel size ≤ 3 mm).

Regarding renal DCE-MRI sequences, experts agreed by consensus that 3D (cartesian or radial) gradient echo-based (GRE) sequences should be used, in combination with a T1-mapping preparatory sequence. Experts commented on using a wide variety of GRE-based sequences, therefore the specific type that should be used was not agreed by consensus.

Consensus recommendations for the sequence parameters of flip angle (FA), repetition time (TR), echo time (TE), and temporal resolution were only agreed upon by experts after defining the previous recommendations described above for renal DCE-MRI planning and sequence parameters. Therefore, final recommendations regarding FA (10 - 13 degrees), TR (shortest available for given FA), TE (shortest available for given FA and TR), and temporal resolution (shortest available, ideally ≤ 3) are contingent upon the user adhering to all other recommendations when implementing a renal DCE-MRI protocol.

Based on feedback from some of the experts, a statement was proposed that the duration of the dynamic scan should be up to 7 minutes, which reached consensus amongst the panel. However, some experts commented that it is not necessary to impose a limit on the scanning duration, and that longer scan times may be appropriate depending on the case, e.g., if there is an obstruction present.

In the final recommendations, experts also agreed by consensus that motion correction should be performed using post-processing image registration. However, recommendations regarding specific registration methods were not agreed by consensus.

A recommendation which did not reach consensus stated that the acquired voxels must be isotropic. However, experts commented that while they believed isotropic voxels should ideally be acquired, this may not always be possible due to temporal resolution requirements and maintaining SNR.

Experts reported using a variety of reconstruction methods (vendor programmes, offline, in-house), however a consensus statement was not proposed for this due to the variety of methods used and lack of comparative studies of existing reconstruction methods.

#### Recommendations regarding renal DCE-MRI modelling and analysis (n = 3)

Regarding arterial input function (AIF) selection, experts agreed by consensus that different approaches should be used depending on the quality of the data. When the data is of good quality, an abdominal aorta AIF should be used, and when the aortic AIF is not of good quality, the data should either be excluded from analysis, or replaced with a population- or model- based AIF. Specific recommendations regarding how aortic AIF ROIs should be drawn were not agreed upon, however experts reported using a variety of methods (e.g., ‘circular ROI in the aorta’, ‘rectangular’, ‘sagittal aortic blood and centred higher in the aorta to minimise inflow artefacts in the abdominal aorta’, ‘automated segmentation selecting a percentage of maximum enhanced voxels’, ‘least-squares fit of the signal to a gamma-variate curve’, ‘near renal arteries (between 5 to 8mm)’).

For the modelling approach, experts agreed by consensus that if using a compartmental modelling approach to analyse renal DCE-MRI data, a two compartment (glomerular plasma and tubular fluid) model for estimating RBF and Glomerular Filtration Rate (GFR) should be used. Experts commented that one compartment models were possible, but not particularly robust. Regarding the use of compartment models with >2 compartments, experts commented that this would be dependent on the quality of the data and the ultimate aim of the measurement. However, experts also commented that >3 compartment models run the risk of overfitting while adding little at the spatial resolution used.

Experts reported using a variety of tools for software post-processing (open access, non-commercial, in-house developed, and commercial). Due to the wide-ranging use, consensus was not found regarding specific software to use. However, experts commented that there is a pressing need for a good open access tool for processing DCE-MRI.

#### Recommendations regarding reporting standards (n = 4)

For recommendations regarding units, experts were asked to provide their responses while taking the following terminology into consideration:

*Perfusion (F_p_) = amount of plasma [mL] delivered per unit of time [min] to a unit of tissue [mL], i.e., [mL/min/mL]*

Based on this terminology, experts came to consensus on the specific units that should be used when reporting Glomerular Filtration Rate (GFR, [mL/min/1.73 m^2^]), Tubular Flow (F_t_, [mL/min/100 mL]), and Plasma Flow (F_p_, [mL/min/100 mL]).

When measuring pixel-based parameters, experts also agreed by consensus that the mean and standard deviation of that measurement should be reported. However, some experts commented that the median and range may be a more appropriate measure if the pixel data are not normally distributed.

## Discussion

By gathering expert opinions and experiences, this paper provides a set of consensus-based technical recommendations to provide a starting point for MRI centres worldwide interested in using renal DCE-MRI.

Initial discussions with the panel revealed that experts applied a mixture of semi-automated, vendor, and in-house analysis protocols when performing renal DCE-MRI, where solutions are dependent upon the metric. For instance, to measure single-kidney glomerular filtration rate (GFR), a different protocol is typically needed than when measuring cortical perfusion. Different approaches in renal DCE-MRI are also required depending on whether the analysis planned is qualitative or quantitative, or if imaging native or transplant kidneys. This results in methodology being established on a case-by-case basis, generating scenarios where experts may struggle to agree on an optimal approach. This, in combination with safety concerns regarding the risk of developing NSF and gadolinium retention in patients with reduced renal function (<30 mL/min/1.73m^2^), has resulted in a decline in the number of studies using renal DCE-MRI. Consequently, challenges arose in identifying key areas where consensus may exist, creating a lengthier and more complex consensus process than for other renal MRI techniques.

The initial survey round, which was formulated as a set of open- and close- ended questions rather than consensus statements, aided in identifying key areas of focus where consensus appeared likely. This initial round also highlighted areas where consensus did not currently seem feasible and provided the experts with an opportunity to provide further insight as to why reaching consensus in these areas may prove difficult in practice. For example, when asked to consider the use of drugs to ensure excretion (e.g., furosemide) when performing renal DCE-MRI, expert responses were mixed. Some experts reported specific protocols used for drug administration, some reported having no experience, and others advised against the use of drugs for such purposes as ‘response to furosemide is variable’ and ‘may alter multiple aspects of renal function’, and ‘furosemide should only be used if you’re trying to measure the effects of furosemide’. One respondent also expressed concern over the clinical feasibility of using furosemide, considering the additional burden this may pose to the patient in addition to the GBCA.

Furthermore, other technical aspects relating to the MRI acquisition could not definitively be formulated as consensus statements due to varied expert responses and experts expressing a lack of experience. For instance, in quantitative DCE-MRI, B_1_-mapping may be required to correct errors in T_1_ measurements caused by magnetic field inhomogeneities (58). However, only a total of three experts reported using B_0_/B_1_-mapping preparatory sequences prior to renal DCE-MRI. Compressed sensing has also been applied as a technique to increase temporal resolution in DCE-MRI (59), however in the initial survey round, ∼38% experts responded that they used compressed sensing in their renal DCE-MRI procedures, while ∼38% responded that they did not. Of those who responded that they did not, one expert commented that they ‘would do if it were available’. When asked to respond to the statement ‘The acquired voxels must be isotropic’, ∼27% responded in agreement, while ∼40% responded that they disagreed. However, when asked to provide a reason, experts commented that isotropic voxels should ideally be acquired, yet it is often difficult to achieve in practice. This revealed that while certain MRI techniques may be useful in renal DCE-MRI, consensus is not currently possible due to a lack of experience, or difficulty in applying optimal techniques in practice. In some cases, experts also reported a lack of experience as it was only possible for them to work with the equipment that was available to them at their centre of work.

Unlike previous renal MRI consensus projects, due to the difficulties that arose in identifying areas for consensus, more technical parameters relating to the MRI acquisition itself (e.g., FA, TR, TE, slice thickness, spatial and temporal resolution) could not be defined in the initial rounds of the Delphi process. Consensus regarding these parameters was contingent on defining consensus in other aspects of renal DCE-MRI first, as slight variations in objectives could result in alternative approaches being taken.

Furthermore, setting certain parameters may conflict with the possible settings for another parameter. Therefore, specific ranges were not provided in the final consensus recommendations for TR and TE as they should be as short as possible, yet what is available on the scanner for one of these values is determined by the value of another, and also the value of the FA. Consequently, in the final consensus statements, it was recommended that an FA between 10-13 degrees should be set first, followed by TR and then TE.

As with the previous consensus studies, the survey responses and discussions also highlighted a critically unmet need for dedicated, reliable, and automated post-processing software for accurate quantification of renal DCE-MRI imaging biomarkers. While a consensus recommendation was not provided in this work, it is hoped that providing a unified starting point for renal DCE-MRI will foster collaboration, aiding developers to progress in this area. However, it will be important for dedicated renal DCE-MRI protocols to be developed and aligned across vendors which also incorporate these consensus recommendations. This is critical for aiding in development of dedicated software which can be used and recognised universally.

A controversial issue regarding renal DCE-MRI analysis relates to the choice of model which should be applied. An inherent trade-off exists between obtaining accurate results and the risk of overfitting. This poses a particular obstacle for promoting uptake in clinical practice. Statements regarding the use of one-compartment models and >2 compartment models could not be included in the final recommendations due to varied responses and ≥40% abstentions. However, from survey responses, experts expressed a clear preference (100% excluding abstentions) for using a two-compartment model if using a compartmental modelling approach to analyse renal DCE-MRI data.

The recommendation of a lower than standard clinical GBCA dose (in general <0.1 mmol/kg of body weight) for renal DCE-MRI is also of relevance considering safety concerns related to the use of GBCAs. In comments received from experts throughout the surveys, it was noted that the main reason for using a lower than standard clinical dose is primarily related to modelling and analysis accuracy. This indicates that the full standard clinical dose which is prescribed may not be necessary for acquiring renal DCE-MRI. Consequently, a reduced dose is not only a safety compromise, but a technical preference that simultaneously addresses GBCA safety aspects, thereby facilitating broader acceptance among referring nephrologists.

### Expert perspective: Paediatric imaging considerations

Most panel members involved in this study had experience only in adult renal DCE-MRI. Consequently, the finalised consensus statements are recommended for use in adults only. However, renal DCE-MRI is particularly valuable in paediatric populations, where congenital and developmental disorders predominate and early functional assessment may influence long-term renal outcomes. Therefore, key considerations for paediatric renal DCE-MRI compiled by two expert panel members (SK, S Serai) with specialist experience in this field, are discussed below. While these considerations do not constitute formal consensus recommendations, they provide a preliminary framework for paediatric application.

#### Indications and clinical use cases

Common paediatric indications include congenital anomalies of the kidney and urinary tract (CAKUT), hydronephrosis and suspected obstruction, reflux nephropathy, renal dysplasia, and assessment of solitary or transplanted kidneys (60–62). In children, renal DCE-MRI is most often used to support functional assessment when an anatomic abnormality is known or suspected (e.g., hydronephrosis/obstruction or CAKUT), and when differential renal function or single-kidney function may influence management (63–65). In transplant recipients, DCE-MRI may support evaluation of graft perfusion and filtration dynamics in conjunction with structural imaging (60,62).

#### Acquisition challenges specific to children

Compared with adults, paediatric renal DCE-MRI must account for smaller anatomy, higher heart rates, faster contrast transit, and increased susceptibility to motion, necessitating higher temporal resolution and motion-robust acquisition and reconstruction strategies. Radial sampling schemes combined with compressed sensing (66), together with motion-compensated reconstructions have been shown to substantially improve robustness to respiratory and bulk motion in infants and young children, enabling reliable free-breathing renal DCE-MRI (67,68). In neonates and infants, renal MRI is frequently performed using feed-and-wrap techniques without sedation to minimise anaesthesia exposure and associated risks; however, residual bulk motion during natural sleep remains common and requires dedicated retrospective motion correction strategies for both anatomical and functional imaging (67–69).

#### Quantitative analysis and interpretation caveats

Renal DCE-MRI has been prospectively validated in paediatric cohorts, demonstrating good agreement between MRI-derived glomerular filtration rate estimates and nuclear medicine reference standards such as 99mTc-DTPA (70). Paediatric-specific quantitative tools are available (71–74) for renal DCE post-processing. These quantitative paediatric workflows further benefit from automated analysis tools, including deep-learning–based renal segmentation (75) or advanced offline CS image-reconstruction for radial imaging (76). Despite methodological similarities to adult analysis frameworks, interpretation in children must account for age-dependent renal physiology, particularly in infants. Accurate estimation of absolute GFR requires reliable measurement of the arterial input function (AIF); however, when patient-specific AIF estimation is unreliable, population-based AIF models are often used. As existing population AIF models have been derived primarily from adult data, their direct applicability to paediatric imaging is limited, and in such cases relative functional assessments, such as differential renal function, may be preferred over absolute quantification (62).

### Study limitations

A broad range of experts from multiple countries and disciplines contributed to this work, crucial for avoiding bias and minimising health disparities in clinical translation. However, while the final panel size falls within the suggested range of eight to 23 participants for health sciences research(56), it was still of limited size (n = 15). While the size of the panel may be considered to be relatively large in the context of the size of the field, due to the varied backgrounds, not all experts were confident in responding to every aspect of the survey. In an attempt to overcome this issue and to minimise bias, statements with ≥40% abstentions were either rewritten and proposed as new statements in subsequent iterations of the Delphi process, or excluded from the final recommendations. As a result, recommendations could not be provided for some topics which may be considered useful for performing renal DCE-MRI. As with previous renal MRI consensus work, the recommendations are intended to provide a unified starting point for clinicians and researchers from which to further technical development within the field. Therefore, the finalised recommendations are aimed to be reviewed and revised accordingly every 5 years, based on progress in the field. Consequently, certain statements which could not be included in this iteration due to ≥40% abstentions, or a lack of consensus, may be able to be included in the future.

## Conclusion

In summary, this work provides a set of expert, consensus-based technical recommendations for performing renal DCE-MRI, relating to aspects of patient preparation, MRI hardware and acquisition parameters, and data analysis. The aim of these recommendations is to contribute to harmonisation of MRI scan protocols across sites, facilitating clinical translation. This will hopefully provide a starting point for MRI centres worldwide wishing to use renal DCE-MRI and help to harmonise existing research MRI protocols to aid development and ultimately improve renal DCE-MRI output.

## Acknowledgements

This manuscript is based upon work from the COST Action CA16103 Magnetic Resonance Imaging Biomarkers for Chronic Kidney Disease (PARENCHIMA), funded by COST (European Cooperation in Science and Technology). For additional information, please visit www.renalmri.org.

IAM is supported by the National Institute for Health and Care Research (NIHR) Cambridge Biomedical Research Centre (NIHR203312*). The views expressed are those of the author and not necessarily those of the NIHR or the Department of Health and Social Care.

FGZ was supported by the German Federal Ministry of Education and Research (BMBF) under the funding code 01KU2102 and 01KU2504, under the frame of ERA PerMed.

IAD was supported by the Dutch Science Foundation (ZonMW, IAD Veni:09150162210040).

## Declarations

### Data availability

Supplementary material for this study is openly available in Zenodo (DOI: 10.5281/zenodo.19709075), containing:

1. Iterative survey rounds, circulated electronically to the panel via Google Forms
2. Finalised consensus statements
3. Excluded consensus statements

### Author contributions

Conceptualization: All authors; Methodology: ERG, IAD; Formal analysis: ERG, IAD; Investigation: ERG, IAD; Writing - original draft preparation: ERG, IAD, AJM, SK, S Serai, S Sourbron; Writing - review and editing: All authors; Data curation: ERG; Visualization: ERG, IAD; Supervision: IAD

